# Bridging Gaps: Improving access to general practice for and with marginalised patients- “it’s quite joyful for us, it’s really improved our work”

**DOI:** 10.1101/2023.05.24.23290453

**Authors:** Lucy C Potter, Tracey Stone, Julie Swede, BG group, Florrie Connell, Helen Cramer, Helen McGeown, Maria Carvalho, Jeremy Horwood, Gene Feder, Michelle Farr

## Abstract

**Background:** People with severe and multiple disadvantage (SMD-combinations of homelessness, substance misuse, violence, abuse and poor mental health) have high health needs and poor access to primary care.

**Aim:** To explore perceptions and experiences of improving access to general practice for people with SMD in healthcare staff and people with lived experience.

**Design and Setting:** Bridging Gaps is a collaboration between healthcare staff, researchers, women with lived experience of SMD and a charity that supports them in a UK city. We co-produced a project to improve access to general practice for marginalised patients, that was further developed with 3 inner city general practices.

**Method:** We observed six collaborative service improvement meetings at three general practices and conducted documentary analysis of minutes of a further three meetings. We interviewed nine practice staff and four participants with lived experience. Three participants with lived experience and one staff member who supports them participated in a focus group. Data was analysed inductively and deductively using thematic analysis.

**Results:** Enabling motivated general practice staff with time and funding opportunities, galvanised by lived experience involvement, resulted in sustained service changes. These included: care coordinators and patient lists to support access to patients in greater need and an information sharing tool. The process and outcomes improved connections within and between general practices, support organisations and marginalised patients.

**Conclusion:** These co-produced strategies could be locally adapted and evaluated elsewhere. Investing in this different way of working may improve inclusion of marginalised groups, health equity and staff wellbeing.

**How this fits in:** This study builds on previous work showing that continuity of care, being able to develop a trusting relationship and being proactive are of particular importance in providing care to highly marginalised patients(4, 5, 6, 7, 8). This work describes co-produced strategies including using care coordinators, patient lists and an information sharing tool to support access and continuity to patients in greater need, in addition to rich contextual information on how to shift ways of working to achieve this. In addition to a small team focused on marginalised patients, this study supports the literature highlighting the need for a trauma-informed approach throughout the whole practice team. These co-produced strategies could be adapted and piloted in other practices and areas. Investing in this focused way of working may improve inclusion of marginalised groups, health equity and staff wellbeing.

## Introduction

The concept of access in this study has four key aspects-availability (included direct and indirect costs to the patient), utilisation, service relevance and effectiveness, and equity (the extent to which resources are mobilised to reflect need)(9). Severe and multiple disadvantage (SMD) is defined here using the gender-sensitive conceptualisation of experiencing at least two out of four primary domains of disadvantage: homelessness, substance misuse, victim of interpersonal violence and abuse, and poor mental health), see figure 1(11). In England 2.3 million adults (5.2% of the population) experience two or more of these primary domains in a single year(11). The combined and intersecting effect of multiple sources of severe disadvantage carries an extremely high burden of mortality, multi-morbidity and frailty(12, 13, 14). Despite this need, people with SMD encounter significant barriers to accessing primary care and lower enablement (the impact of the encounter on patients’ ability to understand and manage their health problems)(7, 15, 16). People with SMD are more likely to have negative experiences of healthcare, including stigma and discrimination, which can act as a lasting deterrent to help seeking; appointment systems are often incompatible with their help seeking behaviours(7, 17). These patients are highly marginalised and most general practice does not effectively include them(7, 18, 19, 20, 21).

**Figure 1.**
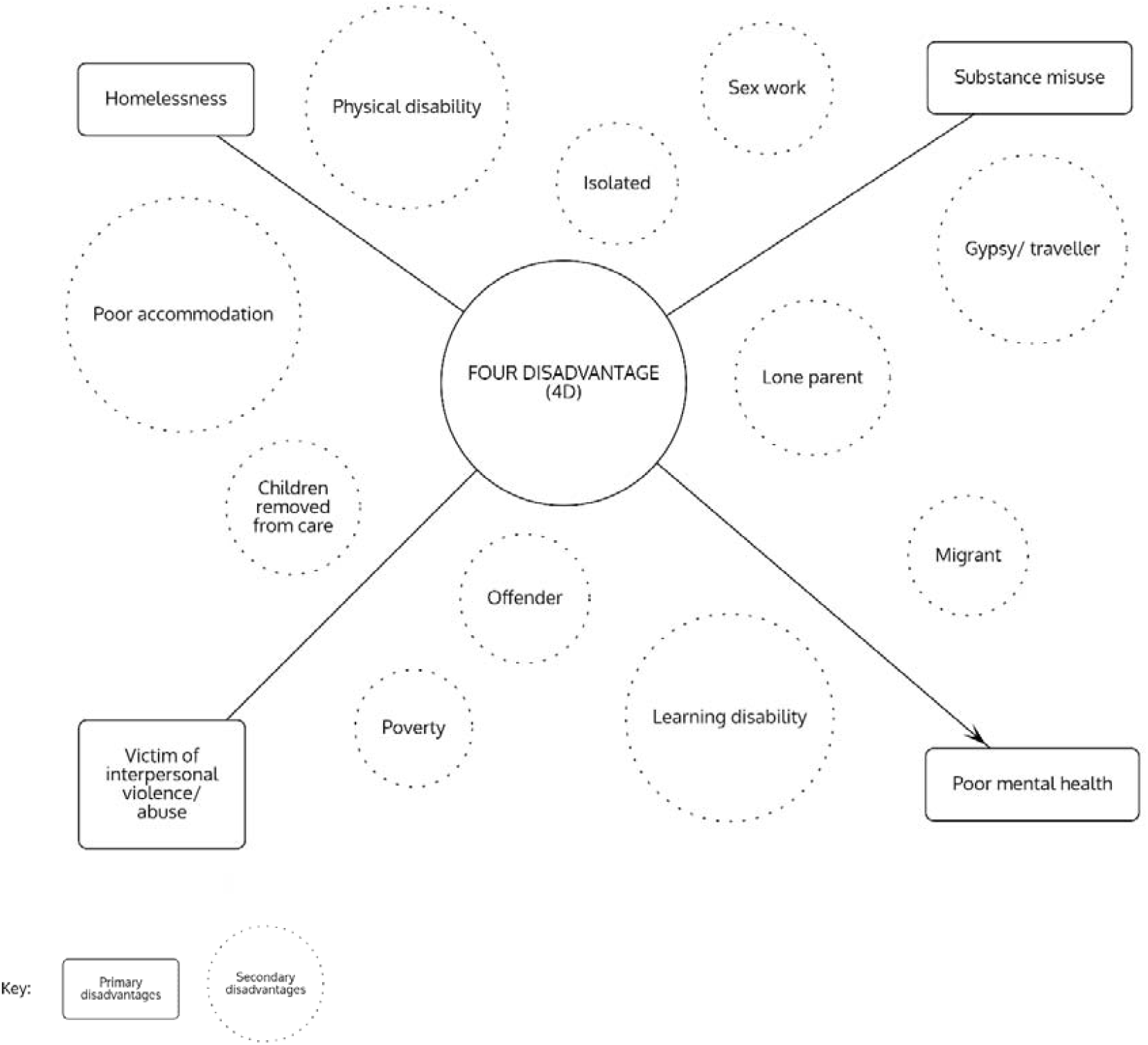
Primary and secondary domains of disadvantage. Reproduced from Sosenko et al 2020(11). Severe and multiple disadvantage (SMD) is defined as experiencing at least two out of four primary domains of disadvantage.

Specialist homeless healthcare centres have emerged in most major cities in the UK to provide primary care to homeless people, but this is only part of the solution. Many people experiencing SMD are not homeless, or may only be homeless temporarily and certain groups such as street sex workers often do not access homeless services due to safety concerns(7). Specialist clinics and outreach are important and useful resources for marginalised communities, but they are often unable to offer the full spectrum of mainstream primary healthcare and often with limited staffing and opening hours. There are also challenges in supporting people to transition from specialist services into mainstream care as crisis situations stabilise and health problems improve (22). Limited outreach or homeless health specialist provision is not enough; there is a need for mainstream primary care to be more inclusive, integrated and accessible to marginalised patients.

General practice is a stretched system in the UK which can be hard for marginalised patients to access, current provision is not proportionate to need(16). Access to care is more than just being registered at a general practice; it requires the ‘human fit’ between the patient and healthcare staff(23). Not being able to provide adequate care to disadvantaged patients contributes to GP stress and burnout(16). Improving the ‘human fit’ between general practice and those most in need is good for patients and staff.

#### Bridging Gaps Project Development

We used a co-production approach, where people with lived experience are involved in decision-making throughout the process(1). The co-production group (Bridging Gaps) was started in May 2019 by LP and women with lived experience of SMD who had been supported by One25 (a Bristol charity which reaches out to some of the city’s most marginalised women). As a GP, author LP had delivered a once a week outreach clinic at the drop-in centre of One25 for two years before the start of the project and had built up a number of trusted relationships. One25 is a women-only safe space and the lived-experience team decided to continue this way. Co-production meetings were held every two weeks and took place in well-known community spaces. Participants were offered shopping vouchers as a thank you for their time. We provide a detailed account of our co-production experience elsewhere(2, 3). Much of Bridging Gaps work focuses on how to make services more trauma-informed(3). Trauma informed care “realises the widespread impact of trauma and understands potential paths for recovery; recognises the signs and symptoms of trauma in clients, families, staff and others involved within the system; and responds by fully integrating knowledge about trauma into policies, procedures and practices, and seeks to actively resist re-traumatisation”(10).

After team building, we contacted general practices identified in areas with higher concentrations of marginalised patients. We held face-to-face sessions with 2 general practices and 1 with the GP training scheme. We held 5 online sessions with general practices during the COVID-19 pandemic. When possible, the group opted to resume face-to-face collaborative work with general practices. We held 3 service improvement meetings at each of 3 GP practices (n=9), this second phase of GP service improvements meetings is the main focus of this paper.

Co-production, where researchers and stakeholders, including professionals and people with lived experience, develop collaborative partnerships, means that marginalised patients can become more equal partners, sharing decision making roles(24). Co-production may result in more implementable interventions and lead to better outcomes(25, 26, 27). The priorities and abilities of marginalised patients and the organisation of health services are poorly aligned and inequalities in access, quality and outcomes in care are worsening(28), creating and exacerbating vulnerabilities(29). Co-production involving people with lived experience and general practice staff offers an opportunity to challenge this discord and bring marginalised patients and health services together.

The aim of this research was to collaborate with people with lived experience, a charity that supports them and general practice staff to co-produce improved access to primary care for people with severe and multiple disadvantage (hereafter referred to as marginalised patients). We sought to articulate the perspectives of marginalised patients, those who support them, and general practice staff participants on their experience of co-producing service improvements and improved access. We focused on the answers to two practical questions:

- What are the key issues and challenges in improving access to primary care for people with marginalised patients?
- What are the potential strategies to improve access to primary care for people with marginalised patients?

## Method

The second phase of the Bridging Gaps project ran between July 2021 and August 2022. Researcher TS joined the project to conduct interviews/ observations with little prior involvement to provide a more independent perspective. Service-improvement meetings at general practices were facilitated by LP and included members of the Bridging Gaps co-production team, a support worker from an organisation that supports them, researchers and selected staff from the general practice. The first meeting at each practice introduced access to general practice for marginalised patients and participants shared their experiences and perspectives, subsequent meetings focused on co-designing plans to improve this. Data collection comprised semi-structured interviews with four lived experience participants; semi-structured interviews with nine GP staff; observations of six of the nine collaborative service improvement meetings by TS (total 12 hours, total 22 participants) with documentary analysis of meeting minutes of the remainder; and a focus group with three lived experience participants and one staff member of an organisation that supports them (by LP and MF).

### Data analysis

Interviews were fully transcribed and coded using QSR NVivo software. Data were analysed (by LP and TS) using reflexive thematic analysis(30). The first two transcripts were coded independently and discussed to explore differences in interpretation and maximise rigour. We used a mix of inductive and deductive (based on project aims) coding and collated the data into themes. Overarching themes were developed by transferring between visual mind maps, narrative text and discussion with the research team.

Several authors had been involved in the co-production team for 2-3 years (LP, MF, FC, MC, HM), lived experience participants who were once marginalised patients over time became colleagues. Reflexivity was essential in harnessing the value of the in-depth involvement of personal experience in this work. Exploring access to general practice for marginalised patients while challenging power relations is complex; reflexive thematic analysis allowed the flexibility to follow an iterative path in order to better understand these. The close involvement of several of the research team including facilitating collaborative meetings meant we were highly familiar with the data and enabled critical engagement throughout.

## Results

30 participants from a range of roles contributed to this phase of the study, including 14 participants in the co-production team and 16 primary care staff, as detailed in Table 1.

**Table 1.** Participant roles

| Role | Number of participants | Category used in quotations |
| --- | --- | --- |
| **Co-production team (Bridging Gaps)** |  |  |
| Patient with lived experience | 8 | Bridging Gaps (BGW) |
| Support worker | 2 | Partner organisation (Ptn) |
| Academic GP | 2 | Researcher (Res) |
| Researcher | 2 | Researcher (Res) |
| **General practice participants** |  |  |
| General practitioner | 6 | General practitioner (GP) |
| Nurse | 2 | Nurse (Nurse) |
| Drug and alcohol worker | 3 | Drug and alcohol (D&A) |
| Care coordinator/ link worker/<br>social prescriber | 5 | Care coordinator (CC) |
| **Total** | **30** |  |

We have distilled the results into three main themes and a summary table (table 2) of the system problems and changes implemented to improve access and care of marginalised patients:

**Table 2.** System problem and change implemented to improve access and care of marginalised patients

| System problem | Change implemented | Illustrative quotes |
| --- | --- | --- |
| General practice and lived experience participants both highlighted that marginalised patients often fall through the gaps of mainstream general practice. | **1. Care coordinator and patient list**<br>Two mainstream practices independently used the opportunity of care coordinator funding to have an individual or team to provide additional support for marginalised patients at their practices. Both practices have continued these roles beyond the project and one is recruiting for a second care coordinator to also do this, because of high levels of need. Participants recognised the value in these individuals maintaining and advocating for a list of marginalised patients. Care coordinators proactively communicate with patients on the list (without 'nagging'), provide continuity and help clinicians to provide appropriate care to them (see Theme 2- Shifting ways of working- provide proactive continuity of care from trusted, supported professionals). | *"The role that (CC1) has is developing into something that's quite essential to the practice for this particular group of people with complex needs, and not only them, but for the staff... it gives us somewhere to put our thinking in terms of how we behave with people with complexity, people we can't get hold of, certain types of vulnerability"-GP2*<br>*"the doctors will send me tasks and say, 'This one needs to be on your radar,'"*-CC1 |
| Mainstream appointment systems are often inaccessible to marginalised patients who often require rapidly responsive, easily accessible routes in to care. | **2. Prioritise flexible access and longer appointments to patients in greater need**<br>One practice dovetailed the care coordinator role with a specialist inclusion health clinic for marginalised patients; in the other the care coordinator had a direct telephone line they provided to patients and priority access to appointments when needed. One practice also had protected appointment slots that could be used by drug and alcohol workers who could use this for one of their clients if needed. | *"I've also got the authority, the clearance, to book appointments in when I need to, whereas the staff, they'd have to say, 'Okay, we haven't got any more for today and the next one's in two weeks,' I can change a slot with the doctor."*-CC1 |
| Several participants reflected the variability in behaviour or prejudice from different staff members and this was a deterrent as they wouldn't know if they would get 'a nice one'. | **3. Trauma-informed approach throughout the whole practice in addition to small team focused on marginalised patients**<br>Both of the mainstream practices we collaborated with on service-improvements requested training for all staff and reflected on the importance of a whole team approach to improving access for marginalised patients.<br>Additionally it was valuable to have a small team of clinicians and care coordinator for patients to have continuity and trust in who they were communicating with. One care coordinator offered patients on her list a direct phone number that would only be answered by her so patients had the security of knowing who they would speak to (see Theme 2- Shifting ways of working- provide proactive continuity of care from trusted, supported professionals). | *"So I think a lot of my barriers was around the way I'd been treated in GP surgeries before. Like I said, not about the GPs, but the receptionists"- BGW2*<br>*"that first meeting was a whole team meeting because I do feel like access to healthcare is very much a whole team issue. It's not just about GPs."- GP1* |
| Lived experience participants raised the need to re-tell their story to multiple staff members as a barrier to accessing general practice. | **4. Personal Snapshot document as a tool to share information with general practice (see Appendix 1)**<br>Participants co-developed a tool to try and mitigate this, a *Personal Snapshot* document that is completed with a support worker and emailed securely to the practice, with an alert on electronic notes to see the document prior to consultation. It is optional, but includes information that the individual may want to share with the practice, whether they would like to be asked about these or not, and best ways of working with them. This uses a trusted connection with a support worker, to help with building a relationship with general practice staff. | *"there's so many services that support people but so few of them are actually able to support people in [general practice]... if you've got one good relationship and somebody can just do that with one person then they've got a much higher chance of being able to communicate what they need"-Ptn2*<br>*"It's really helpful to the GPs to have the background before doing consultations with patients and it gives the patient more confidence and trust in us that we've listened to them and adjusted things to help them."- CC1* |
| Participants raised serious concern about women not feeling safe to access homeless health or drug and alcohol services as the locations are targeted by perpetrators. | **5. Exploring potential changes including healthcare outreach clinics in women's only safe spaces**<br>Participants discussed ideas of collaborating with women's only safe spaces in the city such as women's only hostels or locations that weren't focused on drug use or homelessness to host a regular clinic for an outreach clinician to support marginalised women. Homeless health staff were enthusiastic to support this. As yet funding has not been secured to try this. | *"it's hard to get normality into your life, access accommodation services, when these people (men) are there. The idea of 'drop in' is great but there are all these blokes outside."-BGW2* |

1. Time and funding opportunities enabled motivated individuals and teams to improve general practice with marginalised patients.
2. Participants shifted ways of working: to provide proactive continuity of care from trusted professionals.
3. Improving connections within and between general practice, support organisations and marginalised patients was enjoyable, encouraged empathy and was beneficial to patients and staff.

### 1. Time and funding opportunities enabled motivated individuals and teams to improve general practice with marginalised patients

Lived experience and general practice participants shared experiences of barriers to access for marginalised patients. All participants were motivated to tackle these barriers. Protected, facilitated meeting time with lived experience involvement galvanised professionals who were already interested in improving care for marginalised patients. General practice staff used Primary Care Network care coordinator and Enhanced Access funding to make service-improvements (outlined in Table 2), which have continued beyond this project. This included two practices recruiting a care coordinator to support marginalised patients who struggle to access mainstream general practice, and one practice co-designing an inclusion health clinic. This clinic included support from the care coordinator with appointment booking, reminders and non-clinical support, and longer appointments with one of two GPs experienced in inclusion health. This additional capacity enabled motivated GPs and care coordinators to collaborate on providing better care for marginalised patients.

> *“So it was partly GP13 from the partners and partly the local contract funding that fitted together. So then me and another GP with an interest in inclusion health were given a mandate… to crack on with it and so we did. Then Bridging Gaps was there as a really useful opportunity for people with lived experience who were doing relevant work around improved access for people with experience of trauma. So it felt like a very ideal synergy”- GP5*

When GPs were asked what the hardest part of providing care to marginalised patients was, the most frequent answer was ‘not enough time’. GP participants were conscious of the heightened vulnerability of marginalised patients and referenced really wanting to ‘do a good job’ in providing care for them. Participants had some knowledge of how to do this, but described being limited when simultaneously trying to safely manage high volumes of clinical demand.

> *“if you’re trying to deal with someone with really complex needs in the middle of an absolutely overbooked on-call clinic with 50 calls, and you’re just trying to get through the session safely*…, *you are going to really struggle to provide the empathetic, whole person care that you might want to provide. So structurally, you need to put clinicians in a place where they’ve got the headspace and the opportunity to be kind and trauma-informed and aware of that person’s needs. Otherwise, it’s just not fair on either person*.*”- GP5*

Some participants recognised the potential for overwhelm or burnout in professionals who do not have sufficient capacity for the work, how that can detriment the compassion people feel able to offer and risk staff leaving.

> *“I think that staff are really tired… they’re very kind of stretched and I think that impacts on people’s compassion… staff burnout is I think quite common at the moment”-NURSE1*

One GP who delivered the inclusion health clinic developed during the project in addition to mainstream general practice highlighted how enjoyable it was to have protected time to ‘do a good job’ for patients who really need it.

> *“I think as well as hopefully being good for the patients. It’s quite joyful for us, it’s really improved our work”- GP1*

Another GP who delivered the inclusion health clinic developed during the project in addition to mainstream general practice shared the potential of the inclusion health clinic to improve efficiency and reduce stress elsewhere in the system, as standard clinics are not appropriately timetabled for managing high levels of complexity that could be managed in the inclusion health clinic.

> *“I think, overall, that [colleagues] were supportive of [the inclusion health clinic]… allowing a space for patients who needed more time where it was really stressful to try and do that within a general clinic with 15 patients. So hopefully, it offloaded some of the stress*.*” -GP5*

### 2. Participants shifted ways of working: to provide proactive continuity of care from trusted professionals

Several lived experience participants described negative experiences, including feeling judged by or discriminated against by authority figures, that were barriers to accessing care. Examples included having to wait longer than other patients if attending the surgery to collect opiate substitution prescriptions, or having this called out publicly in the waiting room, exacerbating feelings of shame around substance dependence. These feelings mattered and persisted for people; building trust seemed to be key in enabling engagement. Most professional and lived experience participants highly valued continuity as a strategy to achieve this.

> *“Trust is won in small moments, isn’t it, over a period of time? In terms of structural things that facilitate that… continuity really matters. Yeah, it does, particularly for this group of patients*.*”- GP5*
>
> *“[W]hen you are ready, you’re gonna go to that person ‘cause you’ve built that trust along the way. I trust that they’re not gonna railroad me into something”- BGW2*

Some participants referred to the variability of behaviour marginalised patients might receive from different staff members, and therefore it was important to know who they were going to speak to. One care coordinator had a designated phone number which she gave out to patients for direct access to a known contact. Despite fears from colleagues that she would be overwhelmed with calls, this did not happen. The coordinator saw it as a valued tool that helped marginalised patients feel more comfortable to access.

> *“People aren’t over-using it, not at all, but I think it’s just knowing that it’s there, that they feel a lot safer, that they can have that line and it’s only on my desk; nobody else will answer it*.*”- CC1*

Several participants recognised the benefit of building on existing trusted relationships to encourage engagement with other professionals, or help with the practicalities of getting to appointments.

> *“so we actually get that person to feel that it’s okay, if we’re all singing from the same hymn-sheet and saying, ‘You need to go there. Just ask for this person. You’ll be absolutely fine’”- CC1*

Healthcare professionals who deliver enhanced care to marginalised patients highlighted the importance of proactively contacting marginalised patients. They emphasised not being too pushy, and listening to patients’ priorities first before using opportunities to offer assessment or management that might be of the healthcare provider agenda. Proactive, person-centred communication was valued by both professionals and patients, it offered a way to show compassion, and a way to build relationships.

> *“I text people a lot… sometimes I’ll just send a text saying, ‘I hope you’re okay’… especially when people aren’t engaging, just keeping that relationship going and then when you meet up with them, they will sometimes say, ‘oh you know, I like getting your texts, thank you’*.*”-NURSE2*

Lived experience participants encouraged general practice staff to remember ‘the quiet ones’, patients who might not actively ask for or advocate for their needs and are often left behind by general practices that require people to actively seek care and overcome common obstacles like limited appointment slots. This resonated with several practice staff.

> *“don’t forget the people who are traumatised and go quiet and disappear, but at the same time, we mustn’t hound people… allowing the patient autonomy and control where possible”- GP1*

The importance of honest communication and human connection with marginalised patients was highlighted by professional and lived experience participants and enabled progress of patient and professional agendas.

> *“as a service user you walk in and I know when they’re putting on a voice… just be yourself, do you know what I mean?… I think CC1 to me seems, when you meet her in the surgery is how she would talk at home do you know what I mean? It’s just her”- BGW2*

### 3. Improving connections within and between general practices, support organisations and marginalised patients was enjoyable, encouraged empathy and was beneficial to patients and staff

Both the process of co-production and the service-improvements that were developed (see Table 2) initiated or strengthened teamwork and connectedness both within and between general practice teams. Some participants noted the value of relationships and teams within general practice who were focused on delivering better care for marginalised patients. One participant described relief at feeling part of a wider team of practices also motivated to tackle health inequalities.

> *“it’s really kind of encouraging for us to know that there are other GP practices that are kind of passionate about health inclusion that’s you know, refreshing and a relief!”- NURSE1*

Co-producing service-improvements encouraged empathy, human connection and was rewarding to professionals and marginalised patients. Patients and staff were enthusiastic about the benefits of the changes they had made and outcomes for patients, primary care and collaborating organisations.

> *“there is people behind these labels and there is humans and human behaviour and I think we have made the difference. I think we made the difference like in Practice2, I think we made a difference in Area8… I love that and I think that’s a massive thing”- BGW2*

A couple of lived experience participants reflected that their experience of collaborating with general practice staff helped them empathise with them and feel more comfortable and empowered to engage with general practice.

> *“it’s actually kind of broken down my own barriers towards GPs… it’s made me a little bit more confident to speak up and sort of put my view across and know that I’m able to do that; this is just another human being I’m talking to” -BGW1*

General practice participants highlighted the value of meaningful collaboration between those with lived experience and providers in improving contextual understanding. Collaboration improved empathy, challenged assumptions, enabled those with lived experience to feel listened to, and increased the relevance of services to the local population. This new way of working had a different balance of power which general practice participants found insightful and constructive.

> *“when you change the power dynamic and have a meeting like that, where we’re all on an equal footing, I often find that you have unexpected insights… they’ve challenged the way that we’ve thought about things and given us a fresh way of thinking… it’s improved things for us as doctors as well as for our patients” -GP1*

## Discussion

This study presents an example of using highly collaborative and inclusive methods to develop sustained service changes to improve access to general practice for and with highly marginalised patients. The service changes were achieved by enabling and galvanizing motivated individuals and teams with protected time, funding opportunities and the involvement of patients with lived experience. Co-produced service improvements (see Table 2) included:

- using care coordinators to hold and advocate for a marginalised patient list
- prioritising flexible access and longer appointments to patients in greater need
- promoting a trauma-informed approach throughout the whole practice team in addition to a small team of clinicians and care coordinator focused on supporting and providing continuity to marginalised patients
- the development of a Personal Snapshot document as a tool to share information with the general practice.

With protected time and proactive support from care coordinators, GPs enjoyed ‘doing a good job’ for patients who really needed it. The process and outcomes of this work improved connections within and between general practices, support organisations and marginalised patients which was beneficial to all.

### Strengths and limitations

A key strength of this work is the significant involvement of patients with lived experience and those who work closely with them throughout the study. This provided a refreshing and connecting experience for both general practice and lived experience participants and has helped galvanise change focused on what is important to those it is intended to help.

Men were not included in the lived experience co-production team (this was the decision of the team and organisation that supports them). The project was considering how to improve access to all marginalised patients to general practice and the service changes weren’t restricted by gender. The individuals involved cannot represent the full diversity of opinion of marginalised patients. Marginalisation is more damaging to women than men(14) and women who have experienced street sex work/ prostitution are often the least well heard of the inclusion health groups(5), prioritising their involvement and safety took priority. Gender-sensitivity is a vital component of trauma-informed care(10), and seems to be particularly so in SMD.

## Comparison with existing literature

There is currently insufficient systematic review evidence to make clear recommendations on how to improve access to primary care(9). This study is in keeping with other work showing that proactive and continuity of care, and being able to build a trusting relationship are of particular importance in providing care to marginalised patients(4, 5, 6, 7, 8), but furthers this buy demonstrating how these can be achieved in practice alongside other service improvements to improve access for and with marginalised patients.

In the UK, GPs experience high stress levels, particularly in having ‘insufficient time to do the job’ and large numbers are leaving or considering leaving the profession(31). The increased burden of ill health and multimorbidity in socioeconomically deprived areas and fewer GPs per head of need-adjusted population in deprived than in affluent areas, results in high demands on primary care and increased GP stress(16, 32). There is a risk of moral distress in healthcare: the experience of knowing the right thing to do while being in a situation in which it is nearly impossible to do it(33). This study outlines strategies that can offer hope: professional participants who had additional resources and time to better care for patients who really needed it were enthusiastic about the process and outcomes for both patient care and the staff team.

With limited resources, there is an imperative to make decisions on the principles of proportionate universalism, that health actions must be with a scale and intensity that is proportionate to the level of disadvantage(34). The service-improvements co-produced here represent positive selectivism, where targeted approaches (that sit alongside universal services) are used to cater for specific needs(35). The positive selectivism of marginalised patients was managed by small teams in the practices, with the care coordinator holding the patient list and providing proactive connection and advocacy, working alongside a small number of GPs who were experienced and committed to improving care for marginalised patients. This technique, working particularly with those who are most committed to make changes for patients, fits with social movement and healthcare improvement literature(36). Another study has highlighted the opportunity of support roles such as care coordinators, in facilitating timely access to care and embedding relationship-based care into and across routine general practice(37). There are similarities with efforts to implement reasonable adjustments for people with intellectual disabilities and/or autism in healthcare(38) and the potential for shared learning in improving inclusion across marginalised groups.

In addition to small team focus, all participating practices raised the need for an empathetic trauma-informed approach throughout the whole practice team. Marginalised patients need to feel safe that any staff member they encounter will treat them respectfully. The experiences shared by participants and the fact that one care coordinator saw the need for a direct phone line that nobody else would answer for patients to feel safe to call suggests the experience of judgement, stigma and prejudice are still present in health care interactions. This is consistent with other studies(39, 40) but having only one trusted staff member clearly is a precarious connection. While the influence of societal prejudice against addictions, mental ill-health and homelessness alongside other potentially intersecting biases are undoubtedly challenging to tackle, there is evidence that staff training can improve trauma-informed knowledge, attitudes and behaviours(41).

## Implications for research and/or practice

Healthcare access has been recently described as the ‘human fit’ between the needs and abilities of people in the population and the abilities and capacity of people in the healthcare workforce, in the context of particular societal conditions and organisational structures and processes(23). Our findings support and extend this by highlighting the structural, personal and relational elements that support improved access for marginalised patients. This highly inclusive work helps us move towards a better human fit and relationship between general practice and marginalised patients.

As occurred in our study, co-production can bring fresh thinking to complex problems(25, 26, 27). Some general practices or networks may wish to try similar highly inclusive methods of collaborating with local support organisations and people with lived experience to develop context-specific service improvements. The investment in doing this properly should not be under-estimated: Bridging Gaps has been a four-year project in the making that started from already established trusted relationships(2, 3). A more feasible strategy would be to use or adapt the strategies we have developed here, outlined in table 2, with important contextual understanding contained in the results and discussion. We would encourage others to strive for a level of meaningful engagement with marginalised patients, perhaps in collaboration with a local support organisation, as part of adapting these interventions locally.

Further research is needed on understanding how to achieve trauma-informed care across the whole practice, what processes and outcomes are important in improving access and care of marginalised patients in general practice, and a more nuanced and rigorous understanding of what works for whom and in what circumstances.

## Conclusion

Building on the foundation of a highly inclusive co-production project, enabling motivated general practice staff with time and funding opportunities, our intervention resulted in service changes to improve access to general practice for marginalised patients. Practices created small focused teams in addition to striving for a whole team trauma-informed empathetic approach. Professional participants found joy in doing ‘a good job’ for patients who really need it and that prioritising continuity for patients who really need it may ease pressure on other general practice clinics. These co-produced strategies could be locally adapted and evaluated elsewhere. Investing in this different way of working may improve inclusion of marginalised groups, health equity and staff wellbeing.

## Supporting information

Appendix 1- Personal Snapshot

## Data Availability

All data produced in the present study are available upon reasonable request to the authors

## Ethics approval

This research was granted approval by the University of Bristol Faculty of Health Sciences Research Ethics Committee, references 93802 and 110882. Health Research Authority approval REC reference: 21/HRA/5057.

## Acknowledgements

We would like to thank all general practice, support staff and lived experience participants for their contributions to the project and efforts to improve access to general practice. Thanks also to Lesley Wye for her expertise and encouragement in setting up and supporting the early years of Bridging Gaps. Bridging Gaps received funding through the Q Exchange by the Health Foundation and NHS England and NHS Improvement and National Institute for Health and Care Research (NIHR); Research Capability Funding through the NHS Bristol, North Somerset and South Gloucestershire CCG grant number RCF20/21-1LP; the NIHR School for Primary Care Research grant number 465; the National Institute for Health and Care Research (NIHR) Applied Research Collaboration West (NIHR ARC West) and the National Institute for Health and Care Research Bristol Biomedical Research Centre. LP was supported by the Wellcome Trust grant number 23501/Z/21/Z. GF’s salary was supported by the UK Prevention Research Partnership (Violence, Health and Society; MR-VO49879/1), an initiative funded by UK Research and Innovation Councils, the Department of Health and Social Care (England) and the UK devolved administrations, and leading health research charities. The views expressed in this article are those of the authors and not necessarily those of the NIHR, the Wellcome Trust or the Department of Health and Social Care.

