## Appendix 1- Personal Snapshot for "Bridging Gaps: Improving access to general practice for and with marginalised patients- “it’s quite joyful for us, it’s really improved our work”"

**Personal Snapshot:** Sharing information with my GP Surgery

**This form is to share information with your GP surgery so they can understand you better and provide the best care for you.** It has been developed by Bridging Gaps, a team of people with lived experience of complex needs working with GP surgeries to improve access to trauma-informed healthcare. All questions are optional. This information should be sent by secure email to the GP surgery where it will be stored securely on your medical notes. You may also want to request an appointment to discuss this, and you can have a support worker come with you if you would like.

**Date completed:**

**Patient name:**

**Date of birth:**

**Address:**

**Phone number:**

**The best way to contact me is:**
**If you can't get hold of me I'm happy for you to contact (name and phone number):**

**What I want you to know about me:**

**I do/do not want to be asked about these things:**

**What's going well?**

**What I need help with:**

**The best ways to work with me are:**

**Anything else for us to know that is important to you:**

Thank you for completing this form, we know it can be difficult. If you would like to update this information at any time you can do so by sending another one. If you would like to feedback on this or are interested in the Bridging Gaps project, please contact

**Guidance for organisations on filling out a Personal Snapshot**

This form is for patients who struggle to access and engage with GP surgeries, and should be used as an opportunity to share information that they feel is important, so the GP surgery can better understand their needs and support them. It has been developed by Bridging Gaps, a team of people with lived experience of complex needs, working with GP surgeries to improve access to trauma-informed healthcare.

Patients who have experienced multiple traumas often struggle to communicate their needs within the healthcare setting. **We advise that this form is filled out with the support of someone who is already known to and trusted by the person, and that they feel in control of what is being shared.**

The information provided on this form could include a patient's health priorities or stating the things they don't want to talk about - or it might be a chance to give context to the care they need. Many patients will have coping strategies that should be highlighted so that healthcare professionals are able to better support them in difficult situations. **All questions are optional.** This information should be sent by secure email to the GP surgery where it will be stored securely on their medical notes.

**How to fill out this form:**

- Find a private space where you will be able to work undisturbed and make sure that they are comfortable
- Explain the purpose of the form, and that it will be sent to their GP and stored on their medical records
- Check that they feel safe discussing this information with you
- Let them know that you can stop and come back to it if needs be
- Spend some time talking through the patient's current healthcare needs and how these might be supported
- Think about what good experiences you have of accessing healthcare
- Try to think of ways they could have more positive experiences of accessing appropriate healthcare
- Write down the key points that are relevant in the form and check they are happy with what you have written
- Let them know they will be able to change or update this record by contacting their GP surgery or submitting a new form
- You may want to support the person to request an appointment to discuss their health needs, potentially offering to attend with them to help them make this step.

Thank you for completing this form, we know it can be difficult. If you would like to update this information at any time you can do so by sending another one. If you would like to feedback on this or are interested in the Bridging Gaps project, please contact

### Guidance for General Practices receiving a Personal Snapshot

This form is for patients who struggle to access and engage with GP surgeries, and should be used as an opportunity to share information that they feel is important so the GP surgery can better understand their needs and support them. It has been developed by Bridging Gaps, a team of people with lived experience of complex needs, working with GP surgeries to improve access to trauma-informed healthcare.

We would suggest filing this document to the patients notes and setting an alert on the patients record:

**'Please see Personal Snapshot document dated xx/xx/xx before consulting.'**

This document should then be sent to the Care Coordinator (or equivalent) for the practice to consider whether any further action needed. Patients using a Personal Snapshot may request to remove or replace the document.

We would suggest it is safest to hide the document from the online record, in case of domestic abuse/ safeguarding concerns see https://irisi.org/changes-online-medical-records-gp/ for further guidance on this).

**[Supporting Organisation] and [Name of Surgery]**
**Service User Consent Form - Use of my Personal Information**
*If you would like this form in an alternative format, such as large print, please let us know*

This form is to get your permission for [**Supporting Organisation]** to collect and share information about you that you would like to share with your GP surgery. We will only share it however you approve below. If you'd like, we can also store this information for you so that you can easily read it again at a later date. You can ask to review or change your consent at any time.

Please let us know which permissions you would like to give:

| **Organisation** | **Consent** |
|---|---|
| *Stored* by<br>**[Support Organisation]** | Yes ☐ No ☐ |
| *Shared with & Stored by*<br>my GP surgery | Yes ☐ No ☐ |

**Print name:**

**Signature:**

**Date:**

Thank you for completing this form, we know it can be difficult. If you would like to update this information at any time you can do so by sending another one. If you would like to feedback on this or are interested in the Bridging Gaps project, please contact
